# Multicenter Clinical Comparison of the 10 Minute AMDI™ Fast PCR Mini Respiratory Panel and the Cepheid Xpert Xpress CoV-2/Flu/RSV *plus*

**DOI:** 10.1101/2025.11.04.25339522

**Authors:** Aravind Srinivasan, Regina Martin, Gregory Allen, Christopher Price, Deon Miller, Shilpi Mittal, Christi Ledo, Estibaliz Alvarado, Yvette Arroyo, Francesca Foltz, Patrick Halle, Jacob Amos Hambalek, Alem Abebe, Yujia Liu, Aiying Sun, Humaira Janwari, Kirk Allen, Hsiang-Wei Lu, Noe Gutierrez, Justin Nako, Brenda Martinez, Patrick Walsh, Nicholas Johnsen, Christopher Ross, Nicholas Casarez, Alyssa Fiore, Liza Perez, David Wright, Sarkis Babikian, John McDonald, Horacio Kido, Steve Kuriyama, Ralph Davis, Tammo Heeren, Brian Miller, Frank Gill, David Okrongly, Regis Peytavi

**Affiliations:** Autonomous Medical Devices Incorporated, Santa Ana, CA; Center for Medical Research LLC, Ocean State Urgent Care, Smithfield, RI; ERA Health Research, Redmond, WA; EmVenio Clinical Research, Riverdale, GA; Care United Research, Forney, TX; EmVenio Clinical Research, Kansas City, KS; Autonomous Medical Devices Incorporated, Menlo Park, CA; RotaPrep Inc., Tustin, CA

**Keywords:** CLIA-waived, Influenza, molecular diagnostics, multiplex, point-of-care testing, Respiratory Syncytial Virus, RT PCR, SARS-CoV-2

## Abstract

RT-PCR is the gold standard to determine the etiology of respiratory viral infections caused by SARS-CoV-2, Influenza A, Influenza B, and Respiratory Syncytial Virus. Multiplex testing panels can test for infections or co-infections with these viruses at the point of care. Treatment using antiviral drugs and/or monoclonal antibodies and public health measures using vaccines and specific infection prevention practices depend upon accurate diagnoses. The AMDI Fast PCR Mini Respiratory Panel is a 10-minute RT-PCR test for use at the point of care to detect these viruses from a single anterior nasal swab. It is run on the Fast PCR Instrument with integrated cloud connectivity for data management. The clinical performance of the Fast PCR Mini Respiratory Panel test was established in a multi-center clinical study of 1906 participants using the Cepheid Xpert Xpress CoV-2/Flu/RSV *plus* test as the comparator. The study population included 242 Flu A, 77 Flu B, 71 RSV, and 87 SC2 positive samples and had an overall percent agreement of 97.2% (CI 96.4-97.9%), 99.7% (CI 99.4-99.9%), 99.3% (CI 98.8-99.6%) and 99.3% (CI 98.8-99.6%), respectively. Out of the total 85 (4.4%) discrepant results, 61 (72%) were associated with a low viral load. The AMDI Fast PCR Mini Respiratory Panel has excellent clinical performance, providing clinically useful information at the point of care in less than 10 minutes.

## Introduction

Patients with respiratory illness due to different viruses may present with similar symptoms such as fever, sore throat, runny nose, and myalgia. Even experienced clinicians are unable to determine the etiology of acute respiratory illnesses due to infections with common respiratory viruses such as Influenza A (Flu A), Influenza B (Flu B), Respiratory Syncytial Virus (RSV), and SARS-CoV-2 (SC2). Further, several of these viruses share similar seasonal patterns, increasing the likelihood of co-infections (1). The specific viral etiology of a respiratory infection may inform treatment decisions for some patients using specific antiviral medications and/or monoclonal antibody therapies (2,3). Specific diagnoses are also needed for public health surveillance and prevention measures. A specific viral diagnosis guides infection preventionists to implement measures needed to prevent transmission to vulnerable individuals (4).

Recent guidelines issued by the Infectious Diseases Society of America (IDSA) advise the use of molecular diagnostic tests to determine the underlying cause of these infections (5). However, early in the COVID-19 pandemic, due to the shortage of molecular tests, the initial management of suspected cases used burdensome interventions and isolation procedures until results were available from molecular diagnostic tests run in central labs. The US Government’s emergency use authorization in response to the global COVID-19 pandemic accelerated the development of tests with multiplex capability to detect several of these respiratory viruses from a single nasal swab sample at the point of care. These multiplex tests provide objective determination of the virus involved with that patient, allowing the clinician to be unbiased by the seasonal patterns or the local infection patterns of the included viruses. Multiplex tests also identify coinfections without the need for an additional sample.

Anti-viral treatments are more effective and, in some cases, only approved for use when started early after the onset of symptoms. A PCR based test has at least 100x greater sensitivity than an antigen test (6,7) and is more likely to correctly identify a viral infection early, when it is most effectively treated. A timely diagnosis also limits exposure risk to healthcare workers and vulnerable contacts (8). Timely access and shorter turnaround time to obtain specific pathogen results make multiplex molecular point of care attractive in healthcare workflows. Many different point-of-care systems are now widely used in outpatient clinics and urgent care facilities for specific diagnosis of upper respiratory infections. While most of these tests have very good sensitivity and specificity, they vary in the sample preparation and total turnaround time taking anywhere from ∼20 minutes (Roche Cobas Liat^TM^) to ∼36 minutes (Cepheid Xpert Xpress^TM^) to provide results after introduction of the sample (9)

The AMDI™ Fast PCR System consists of the Fast PCR Instrument and the Fast PCR Mini Respiratory Panel (Fast PCR MRP) test disc. The Fast PCR MRP uses multiple reverse transcriptase polymerase chain reaction (RT-PCR) assays to detect the presence of Flu A, Flu B, RSV, and SC2 in an anterior nasal swab (ANS) specimen. The Fast PCR Instrument automatically processes the Fast PCR MRP test disc after an operator loads the sample and places the test into the Fast PCR Instrument. Data processing and interfacing with the electronic health record (EHR) is automatically done through a cloud connection.

This paper presents the results from a multicenter clinical study comparing the performance of the Fast PCR Mini Respiratory Panel to an FDA-cleared CLIA-waived comparator test (the Cepheid Xpert Xpress CoV-2/Flu/RSV *plus*).

## Materials and Methods

### Study Design and Clinical Samples

A total of nine (9) point of care clinical testing sites in the United States holding a CLIA Certificate of Waiver were selected to participate in this clinical agreement study. None of the operators had prior training on the use of the Fast PCR System. The installation and operation of the Fast PCR Instrument and testing of all samples from study participants using the Fast PCR MRP were performed by the operators at the clinical sites using only a Quick Reference Guide (QRG). The clinical study sites were either urgent care clinics or adult/pediatric clinics that provide care for outpatients who present with common respiratory symptoms. A total of 28 operators across the nine study sites conducted the testing using Fast PCR MRP system in the study.

Participants that presented with symptoms of respiratory infection were consented and enrolled in the clinical study (see Supplementary Table S1 and Table S2 for complete inclusion/exclusion criteria). Each participant provided anterior nasal swab (ANS) specimens in both the proprietary AMDI Sample Buffer and Universal Viral Transport Media (BD-UVT) (BD Part# 220220) for comparator testing. Since each participant provided two samples, each site alternated the order of swabs collected for consecutive patients to minimize sample collection bias.

A total of 1947 participants that presented with symptoms of upper respiratory infection were prospectively enrolled over a 6-month period (Dec 2024 – May 2025) in an “all-comer” fashion across all clinical sites. Due to the low prevalence of RSV during the enrollment period, prospectively collected samples were augmented with archived ANS sample pairs. A total of 132 archived samples were selected and enrolled in the study using a similar set of Inclusion/Exclusion criteria as prospective participants. The final sample population consisted of 1906 ANS sample pairs. Figure 2 summarizes the total enrollment in the clinical study.

**Figure 1:**
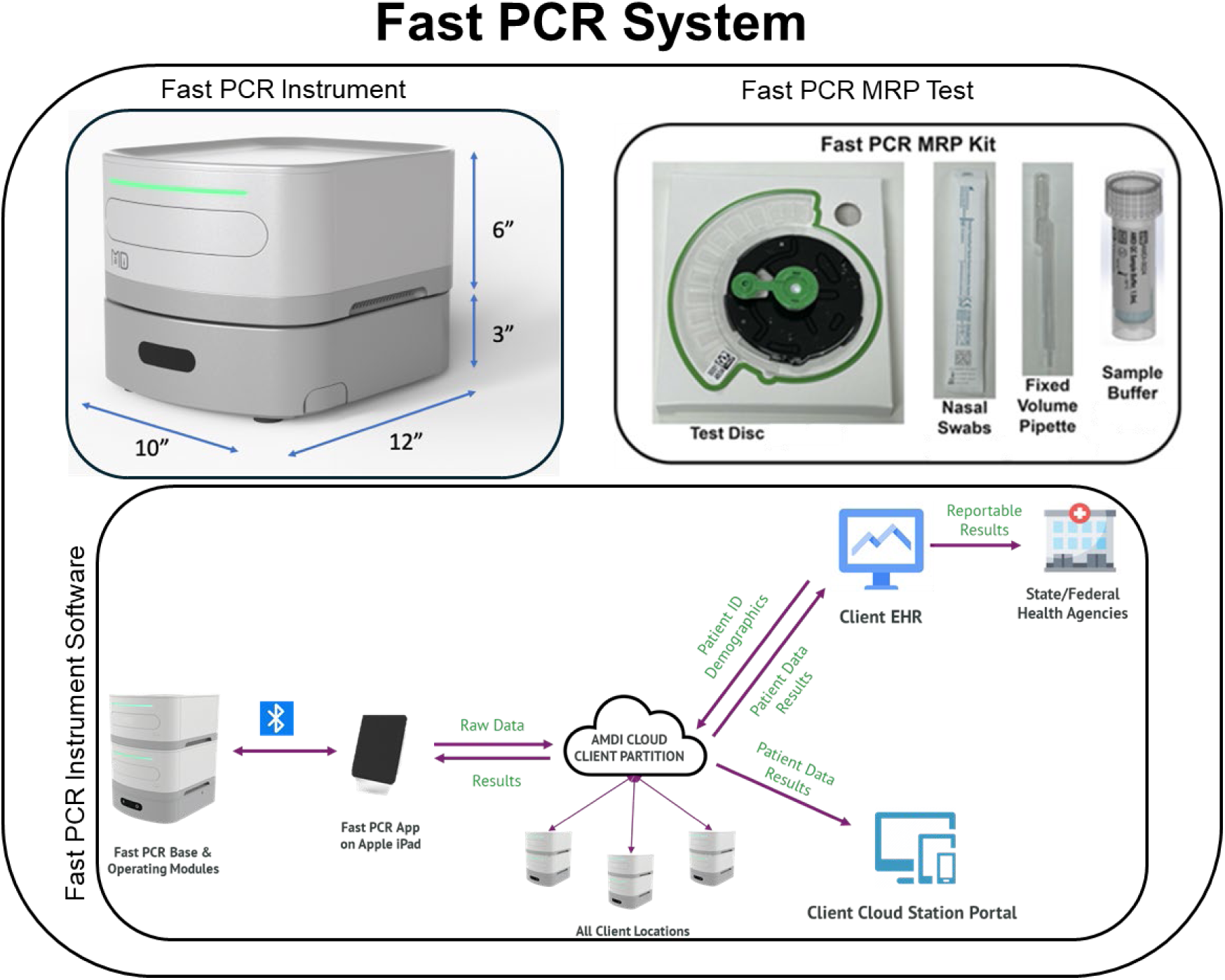
The Fast PCR Instrument consists of the Fast PCR App on an iPad, a Base Station and up to 4 Operating Modules. The Fast PCR MRP Kit contains test discs packed in a test disc holder to facilitate sample addition, fixed-volume transfer pipettes, nasal swabs, and AMDI Sample Buffer. The Fast PCR Instrument provides cloud connectivity required for sample and data processing, results generation, and transmission of results to the patient’s EHR.

**Figure 2:**
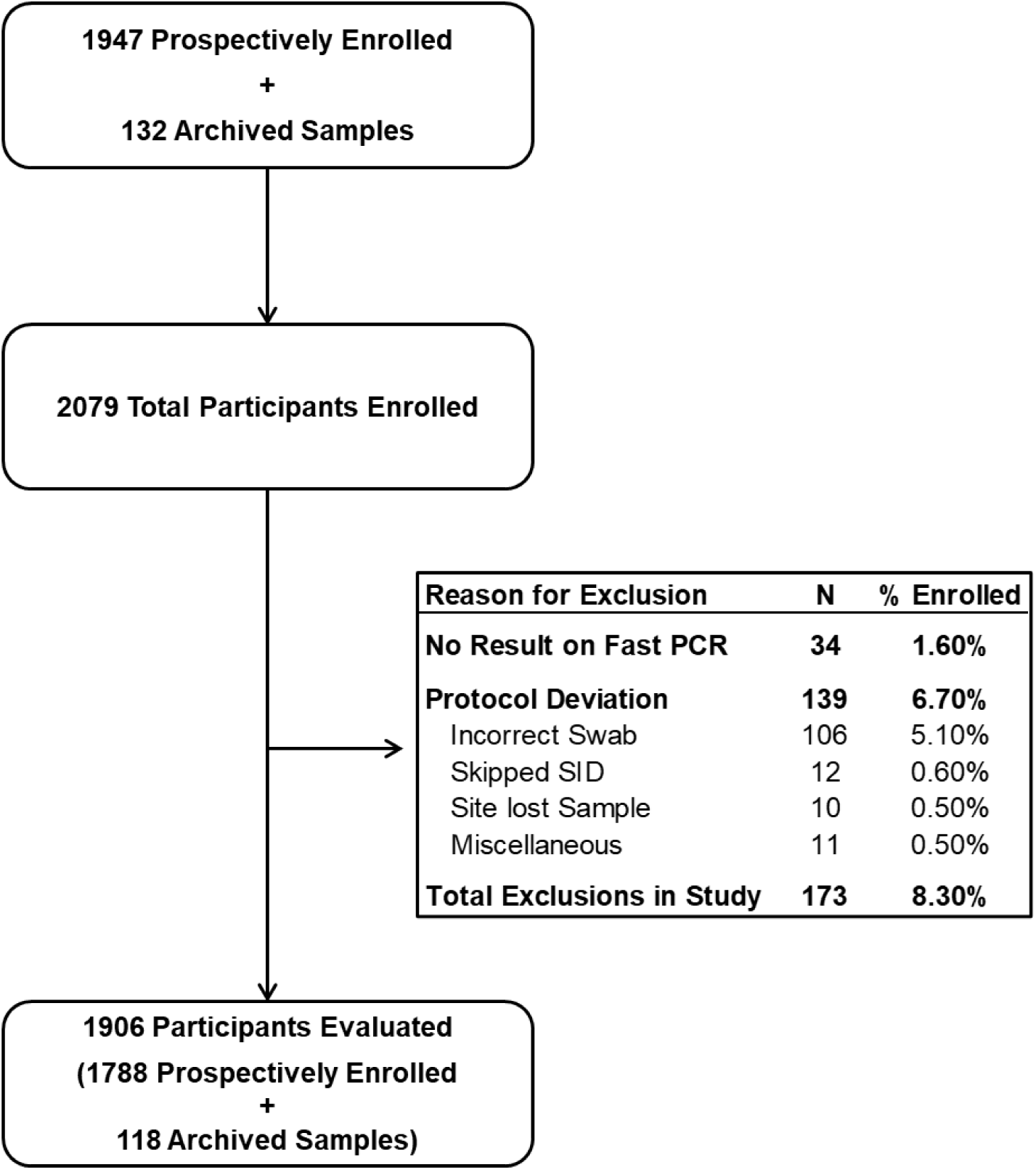
Participants enrolled and evaluated in the clinical agreement study of the AMDI Fast PCR MRP Test

The ANS samples in AMDI Sample Buffer were tested using the Fast PCR Mini Respiratory Panel test at the clinical sites while the ANS samples in BD-UVT were frozen and shipped on dry ice to AMDI (Santa Ana, CA) where they were tested by the comparator method.

### Test Methods

The Fast PCR MRP test is a point of care multiplex RT-PCR test that, in less than 10 minutes and with less than 1 minute hands on time, detects the presence of viral RNA from Flu A/Flu B/RSV/SC2 in an ANS specimen. The test is performed on the Fast PCR Instrument with a single-use disposable disc that integrates sample processing, reverse transcription, amplification and detection in two different optical channels. Result interpretation was done by cloud-based analysis software and the results were returned and displayed on the iPad tablet connected to the instrument.

Comparator testing was performed at AMDI (Santa Ana, CA) on the samples collected in BD-UVT using the Cepheid Xpert Xpress CoV-2/Flu/RSV *plus* test on the Cepheid GeneXpert Xpress instrument according to the manufacturer’s instructions under a CLIA Certificate of Waiver.

### Quality Control

Quality control materials for the Fast PCR System included lyophilized positive and negative external controls and were provided to each clinical site to ensure performance and robustness of the system. The controls were reconstituted with AMDI QC Sample Buffer and run on every Fast PCR Instrument on each day that a clinical sample was tested. For the comparator, quality controls recommended by the manufacturer were run once every week, when a new lot of test kits were received, or when a new instrument was installed. On either system, clinical samples were tested only after quality control procedures were completed and passed.

### Human Subjects

The study protocol was reviewed and approved by institutional review board (Advarra IRB # Pro00082929) before participants were enrolled in the study. In addition, each clinical study site received IRB approval to conduct the study. The study was conducted in accordance with current country and local regulations, and the relevant portions of the International Council on Harmonisation, Good Clinical Practice, (ICH, https://database.ich.org/sites/default/files/ICH_E6%28R3%29_Step4_FinalGuideline_20 25_0106.pdf last accessed 08 Oct 2025 GMT 04:10) and the Declaration of Helsinki 2024 (World Medical Association, https://www.wma.net/policies-post/wma-declaration-of-helsinki/, last accessed 02 Oct 2025 GMT 22:05). Before study-specific procedures were conducted each participant and/or their legal guardian was provided with oral and written/electronic information describing the nature and duration of the study and signed an Informed Consent Form/Assent Form in a language in which they were fluent. Appropriate data protection processes were put in place to protect confidential personal information including participant identities in accordance with the US Health Insurance Portability and Accountability Act of 1996 (HIPAA, USC 45 CFR 160 and 45 CFR 164).

### Data Processing and Statistical Analysis

At the completion of each Fast PCR MRP test, the raw data were automatically transmitted to the AMDI Cloud, processed using the Fast PCR Analysis Algorithm, and the result was transmitted back where it was available on the iPad connected to the Fast PCR Instrument within seconds of test completion. The operators at the clinical site entered the results of the test into an Electronic Data Capture (EDC) database maintained by a third-party vendor.

Similarly, operators that completed the comparator testing of the clinical samples entered the comparator testing data into the EDC database. They did not have access to testing results that were generated at clinical sites using the Fast PCR System until data were locked for statistical analysis.

Positive percent agreement (PPA), negative percent agreement (NPA) and overall percent agreement (OPA) for each target were calculated using 2×2 tables. Wilson’s binomial exact method was used to determine the 95% confidence intervals (CI) of percent agreement. Cohen’s kappa coefficient was used to assess the agreement between the Fast PCR MRP and comparator. Linear regression plots between Fast PCR and comparator Ct values were generated and the Kendall τ non-parametric correlation coefficient was used to evaluate the quantitative relationship between the Ct values generated by the two platforms. The Ct values of discrepant samples were evaluated and reported separately. All statistical analyses were done using JMP Pro 18 (JMP Statistical Discovery LLC, NC).

## Results

### Participant Enrollment and Demographics

Table 1 lists the clinical study sites and number of participants from each site included in the final data analysis. The study population consisted of patients who presented with symptoms of upper respiratory infection such as fever, sore throat, headache, chills, myalgia and runny nose to an outpatient healthcare facility or an adult/pediatric clinic. The study enrolled a total of 2079 participants. After exclusions, the final data set included 1906 participants. The participants were 38.6% male and 61.3% female with a median age of 35 years. The race and ethnic profile of the study population was predominantly white (74.0%) and non-Hispanic (78.6%). Detailed demographic information is summarized in and Tables S1 – S3.

**Table 1:** Clinical study sites, type and number of participants evaluated at each location.

| Site #, State | Type | # Participants |
| --- | --- | --- |
| Clinical Site #1, RI | Urgent Care | 413 |
| Clinical Site #2, WA | Adult Clinic | 315 |
| Clinical Site #3, TX | Adult/Pediatric Clinic | 214 |
| Clinical Site #4, GA | Clinic Attached to Hospital | 165 |
| Clinical Site #5, TX | Urgent Care | 203 |
| Clinical Site #6, KS | Clinic Attached to Hospital | 164 |
| Clinical Site #7, IL | Urgent Care | 73 |
| Clinical Site #8, AL | Urgent Care | 211 |
| Clinical Site #9, WA | Pediatric Clinic | 148 |
| Total |  | 1906 |

**Table 2:** Sex Demographics in the study population.

| Sex | All Participants<br>n (%) | Prospective<br>n (%) | Archived<br>n (%) |
| --- | --- | --- | --- |
| All | 1906 | 1788 (93.8%) | 118 (6.2%) |
| Female | 1169 (61.3%) | 1104 (61.7%) | 65 (55.1%) |
| Male | 736 (38.6%) | 683 (38.2%) | 53 (44.9%) |
| Unknown | 1 (0.1%) | 1 (0.1%) | 0 (0.0%) |

### Clinical Agreement

The study population included a total of 242 Flu A, 77 Flu B, 71 RSV, and 87 SC2 positive samples and 1434 negative samples. The 2×2 tables and the positive percent agreement and negative percent agreement for each analyte are summarized in Table 3a - 3e. Overall, the concordance between Fast PCR MRP and the comparator was excellent with 3 out of the 4 assays (Flu B, RSV and SC2) had a >99% overall agreement. Flu A had an overall agreement of 97.2%. Cohen’s kappa values for the assays ranged between 0.88 – 0.97 indicating near perfect agreement (10). Although neither the Fast PCR Mini Respiratory Panel nor the comparator test are quantitative assays, the regression analysis of the Ct values of samples that were positive on both platforms (Figure 3a - Figure 3d) had slopes of 0.98-1.20 for the four assays and a non-parametric Kendall’s τ correlation of 0.57 – 0.70 (Table 4) indicating a strong linear relationship between the two tests.

**Figure 3a:**
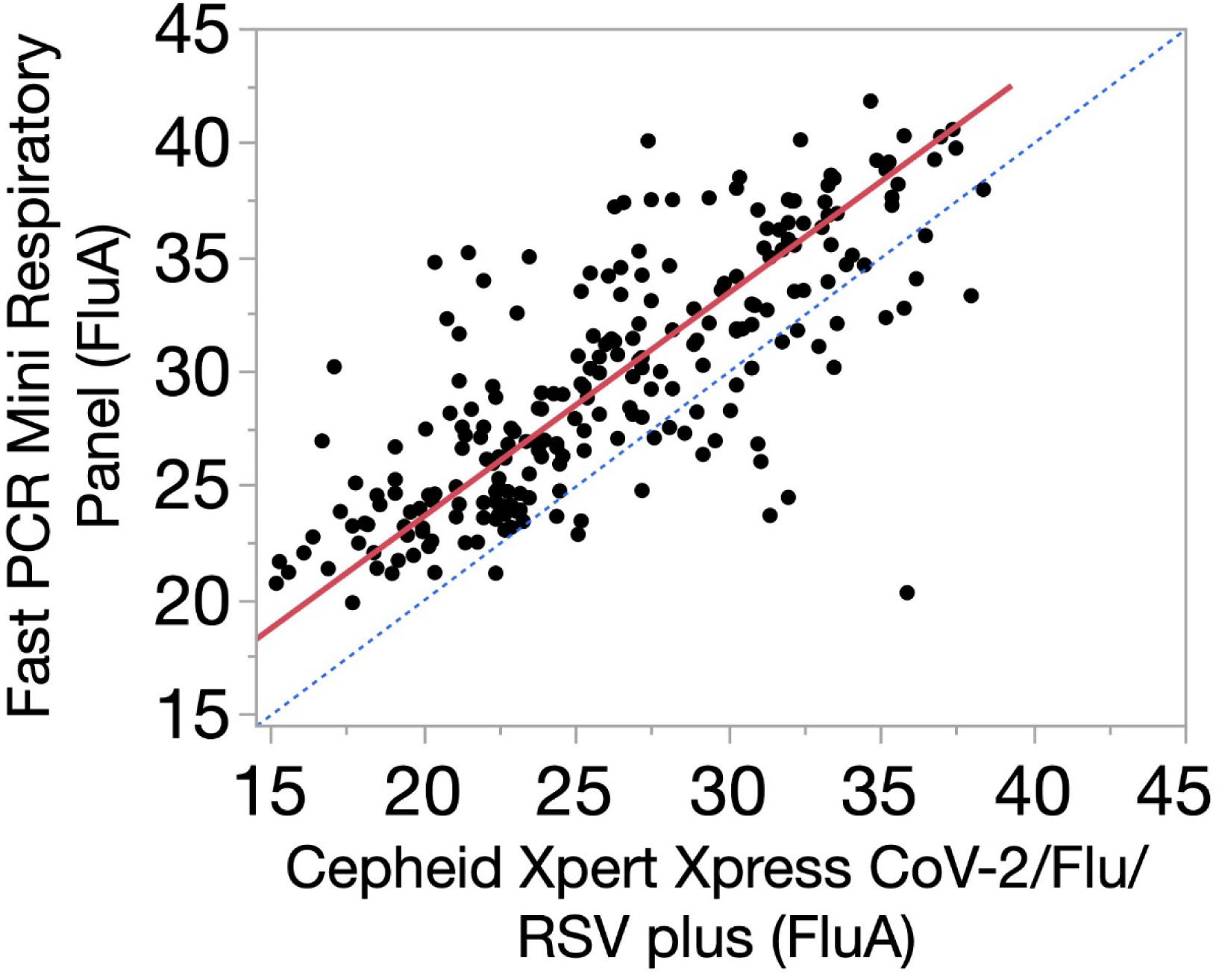
Correlation plot of the Fast PCR MRP Flu A assay cycle threshold (Ct) values with the Flu A1 Ct values from Cepheid Xpert Xpress CoV-2/Flu/RSV *plus* test, N = 236

**Figure 3b:**
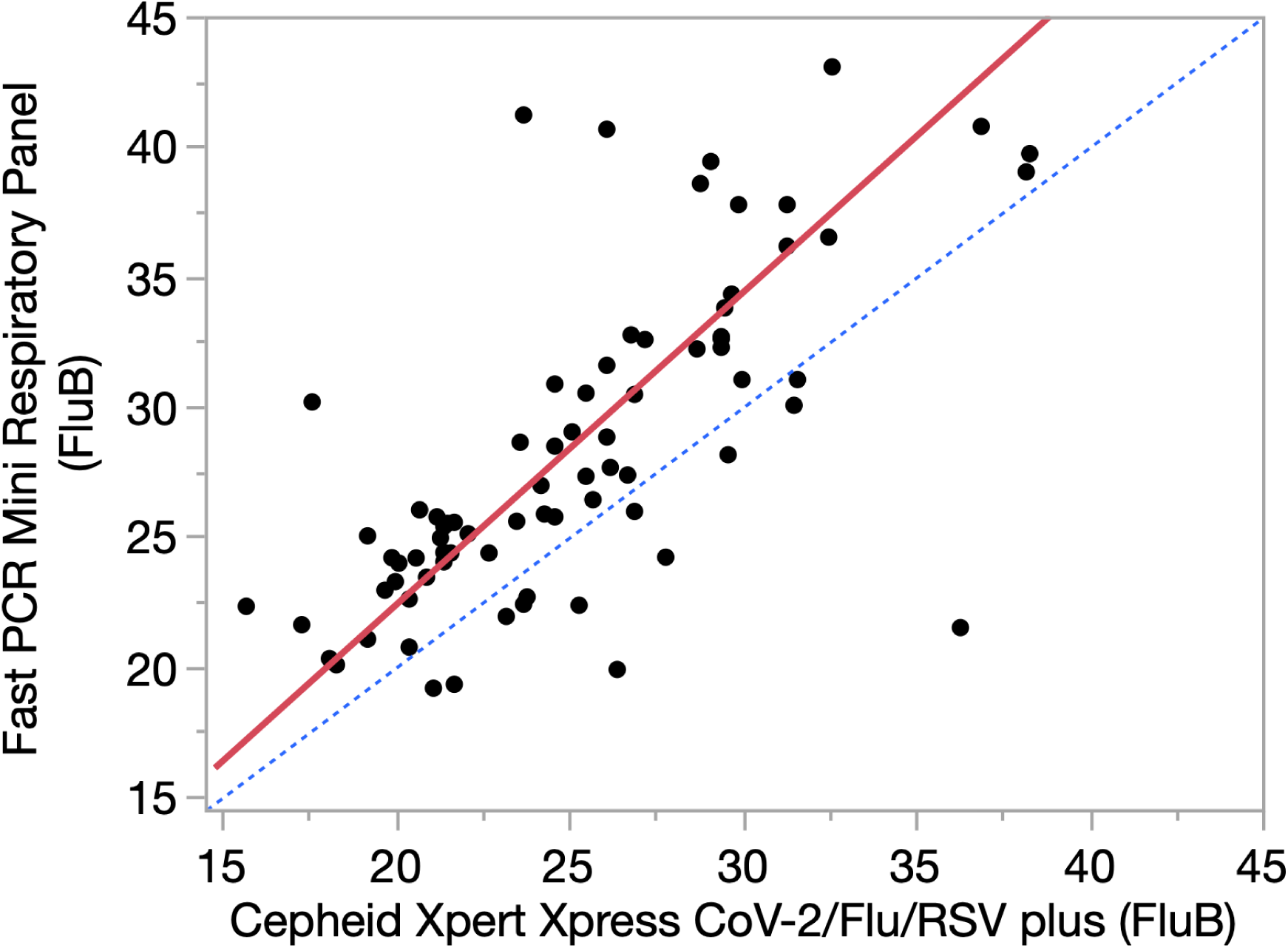
Correlation plot of the Fast PCR MRP Flu B assay cycle threshold (Ct) values with the Flu B Ct values from Cepheid Xpert Xpress CoV-2/Flu/RSV *plus* test, N = 77

**Figure 3c:**
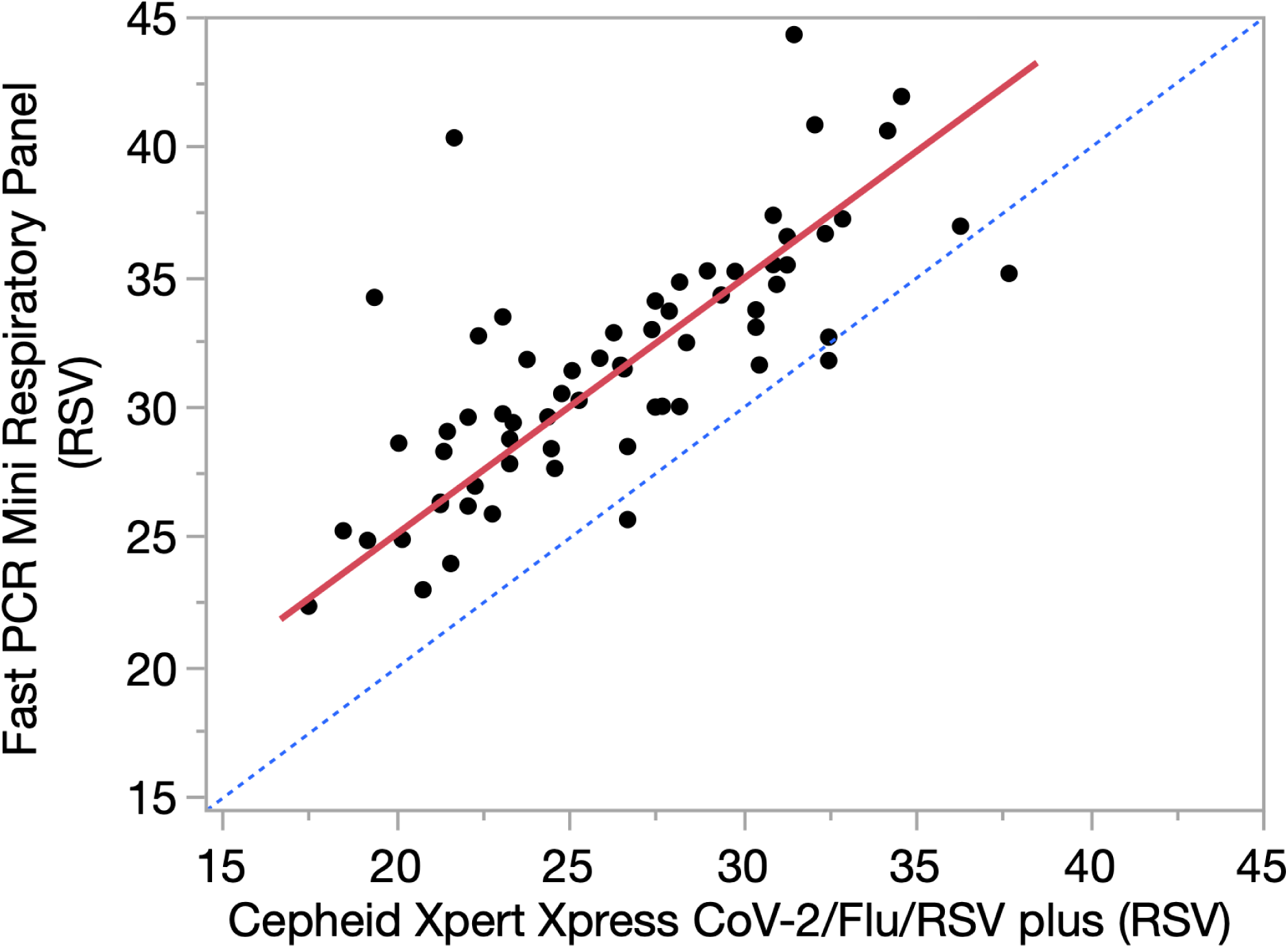
Correlation plot of the Fast PCR MRP RSV assay cycle threshold (Ct) values with the RSV Ct values from Cepheid Xpert Xpress CoV-2/Flu/RSV *plus* test, N = 65

**Figure 3d:**
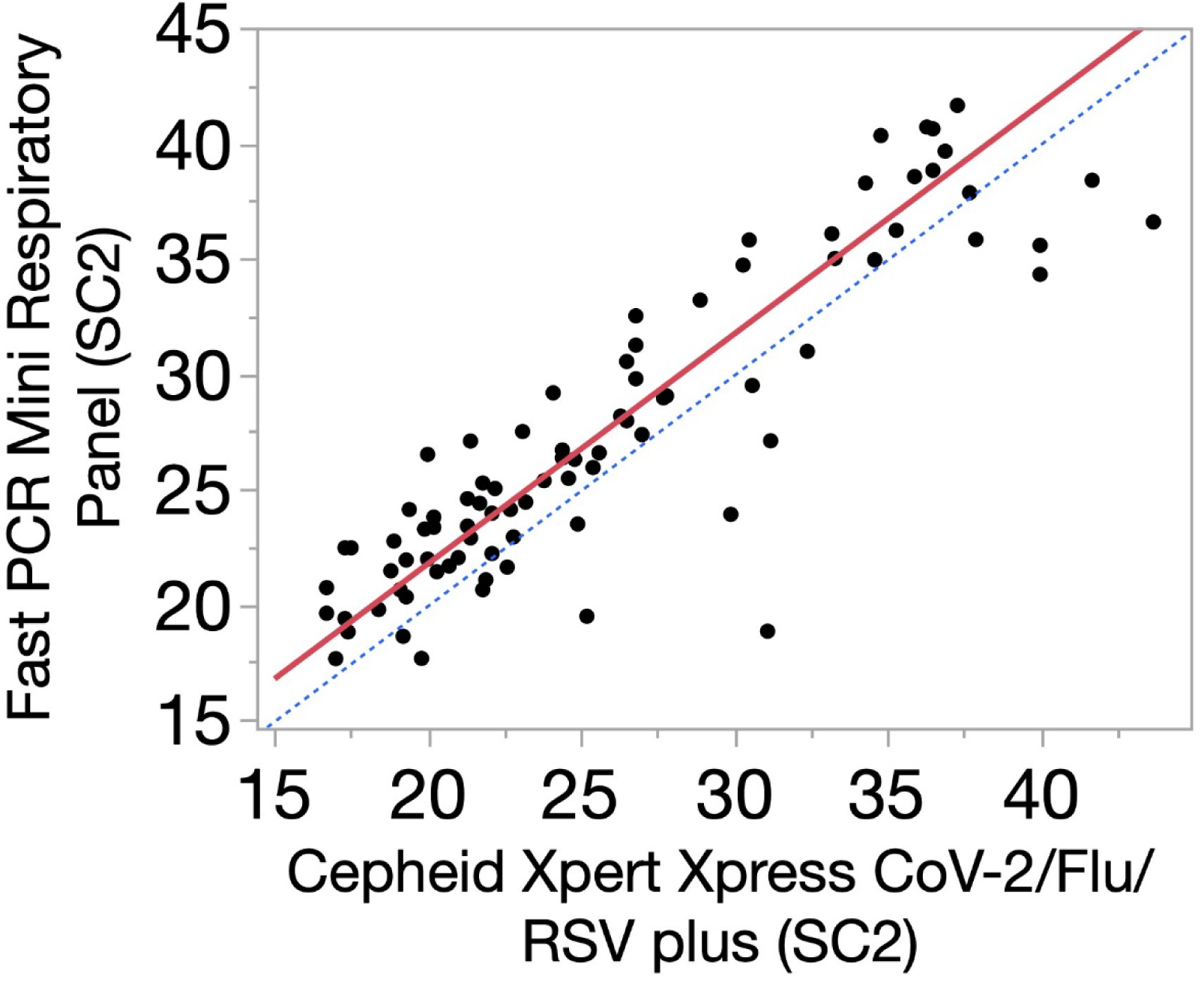
Correlation plot of the Fast PCR MRP SC2 assay cycle threshold (Ct) values with the SARS-CoV-2 Ct values from Cepheid Xpert Xpress CoV-2/Flu/RSV *plus* test, N = 85

**Table 3:** 2×2 tables for a) Flu A b) Flu B, c) RSV, d) SARS-CoV-2 and e) percent clinical agreement of the Fast PCR MRP test with the Cepheid Xpert Xpress CoV-2/Flu/RSV *plus* test.

| Flu A | Fast PCR Pos | Fast PCR Neg | Total |
| --- | --- | --- | --- |
| Comparator Pos | 236 | 6 | 242 |
| Comparator Neg | 47 | 1617 | 1664 |
| Total | 283 | 1623 | 1906 |

b)
| Flu B | Fast PCR Pos | Fast PCR Neg | Total |
| --- | --- | --- | --- |
| Comparator Pos | 77 | 0 | 77 |
| Comparator Neg | 5 | 1824 | 1829 |
| Total | 82 | 1824 | 1906 |

c)
| RSV | Fast PCR Pos | Fast PCR Neg | Total |
| --- | --- | --- | --- |
| Comparator Pos | 66 | 5 | 71 |
| Comparator Neg | 9 | 1826 | 1835 |
| Total | 75 | 1831 | 1906 |

d)
| SARS-CoV-2 | Fast PCR Pos | Fast PCR Neg | Total |
| --- | --- | --- | --- |
| Comparator Pos | 85 | 2 | 87 |
| Comparator Neg | 11 | 1808 | 1819 |
| Total | 96 | 1810 | 1906 |

e)
| Analyte | % PPA<br>(95% CI) | % NPA<br>(95% CI) | % Overall<br>Agreement<br>(95% CI) | Cohen's Kappa<br>(95% CI) |
| --- | --- | --- | --- | --- |
| Flu A | 97.5<br>(94.7-98.9) | 97.2<br>(96.3 – 97.9) | 97.2<br>(96.4 – 97.9) | 0.88<br>(0.85- 0.91) |
| Flu B | 100.0<br>(95.2 – 100) | 99.7<br>(99.4-99.9) | 99.7<br>(99.4 – 99.9) | 0.97<br>(0.94 – 1.0) |
| RSV | 93.0<br>(84.6-97.0) | 99.5<br>(99.1 – 99.7) | 99.3<br>(98.8 – 99.6) | 0.90<br>(0.85 – 0.95) |
| SARS-CoV-2 | 97.7<br>(92.0 – 99.4) | 99.4<br>(98.9-99.7) | 99.3<br>(98.8 – 99.6) | 0.92<br>(0.89 – 0.97) |

**Table 4:** Correlation parameters of Fast PCR MRP test cycle threshold (Ct) with Cepheid Xpert Xpress Cov-2/Flu/RSV *plus* test Ct values.

| Analyte | N | Intercept | Slope | Non-parametric<br>Kendall's $\tau$ Correlation |
| --- | --- | --- | --- | --- |
| Flu A | 236 | 4.03 | 0.98 | 0.60 |
| Flu B | 77 | -1.69 | 1.20 | 0.57 |
| RSV | 65 | 5.41 | 0.98 | 0.61 |
| SARS-CoV-2 | 85 | 1.81 | 1.00 | 0.70 |

### Co-infections

There were 15 samples that were positive for two viral targets (co-infections) when tested with the Fast PCR Mini Respiratory Panel (Supplementary Table S6). Five (5) of these samples were concordant with the comparator for both positive viral targets. The remaining ten (10) samples were identified as co-infections by Fast PCR MRP but not by the comparator method. In nine (9) of these samples, one of the positive targets identified by the Fast PCR MRP was also identified by the comparator. The remaining sample was negative for all targets on the comparator method.

### Discrepant Result

Among the 1906 clinical samples, there were a total of 85 discrepant results (4.4%) observed between the Fast PCR MRP and the comparator (Table 5). Out of these, 61 samples (72%) that were detected as positive by only one of the platforms had Ct value >37.0 indicating a low viral load in the sample tested (10)

**Table 5:** Discrepant results in the clinical agreement study comparing Fast PCR Mini Respiratory Panel test with Cepheid Xpert Xpress CoV-2/Flu/RSV *plus* test.

| Analyte | Discrepant Results with Ct <30 | Discrepant Results with $30 \leq Ct \leq 37$ | Discrepant Results with Ct > 37 | Total Discrepant Results |
| --- | --- | --- | --- | --- |
| Flu A | 5 | 10 | 38 | 53 |
| Flu B | 2 | 0 | 3 | 5 |
| RSV | 3 | 3 | 8 | 14 |
| SC2 | 1 | 0 | 12 | 13 |
| Total | 11 | 13 | 61 | 85 |

## Discussion

Recent reports suggest that, in the current highly immunized population, SARS-CoV-2 viral load appears to peak 3-5 days after onset of symptoms (11). Further, samples containing Omicron variants have been observed to be culture-positive at lower viral loads (12) suggesting that individuals might be infective earlier after onset of symptoms. Due to the low sensitivity of antigen tests, individuals may not reliably test positive on an antigen test for up to 5 days after onset of symptoms, a period when viral load is increasing and the individual is presumably infectious. In the current test-to-treat paradigm for respiratory illnesses, this crucial early window when most antiviral treatments are effective will be missed by relying on rapid antigen tests. The use of molecular diagnostics tests is therefore important for both patient management and for public health practices.

Multiplex molecular diagnostic tests have become the standard means of diagnosing viral respiratory infections due to their overlapping seasonality and possibility of co-infections (13). Antiviral medications and/or monoclonal antibody treatments have been developed for SC2, Influenza, and RSV, raising the value of specific diagnoses of infections due to these viruses. Recommendations from public health agencies and decisions about vaccine use are better informed using data on specific viral infections.

However, molecular diagnostic workflows in central core laboratories are cumbersome and time consuming with routine turn-around times of 24-48 hours. Because early diagnosis of viral etiology is critical for appropriate intervention with specific antivirals, relying on results from core laboratories is impractical. As a result, the optimal testing site for common respiratory infections has moved away from central core laboratories and toward outpatient settings near the point of care where patients with respiratory symptoms first present. In response to the COVID-19 pandemic, several point-of-care systems have been developed and provide frontline healthcare workers with actionable results. These systems all have excellent clinical sensitivity and specificity (14–16) but differ greatly in the total turnaround time.

The AMDI Fast PCR Mini Respiratory Panel is a multiplexed RT-PCR molecular diagnostic test for four common respiratory viruses; Flu A, Flu B, RSV and SC2 from a single anterior nasal swab in CLIA-waived settings. Each MRP test disc has a built-in RNaseP assay that serves as a sample adequacy control providing feedback on potential sampling or sample-loading errors. Data processing is done remotely in the AMDI Cloud and the results are available within seconds both in the patient’s electronic health record and on a dedicated iPad connected to the Fast PCR Instrument. The sample-to-answer time of the Fast PCR MRP test was ∼10 min compared to the ∼36 min for the comparator, a vast improvement over existing platforms to provide an actionable result at the point of care.

In this multicenter clinical study, 1,906 participants were enrolled in an “all-comer” fashion at 9 point-of-care clinics in the United States. The number of participants and their geographic location was selected to minimize population bias. The Fast PCR MRP test displayed excellent qualitative agreement. All the measures of agreement, the PPA, NPA and OPA and the Cohen’s kappa values, showed near-perfect agreement for all four analytes with a widely used FDA-cleared CLIA-waived non-reference benchmark comparator test. Post-study surveys of the Fast PCR System operators (not presented here) documented the ease of use of the system in point-of-care CLIA-waived settings.

The slope of the regression between the cycle threshold values of the Fast PCR MRP and the comparator tests were close to unity for all analytes indicating an excellent quantitative correlation between the two tests. The non-zero intercept for all the assays may be due to differences in the gene targets used by the Fast PCR MRP and the Cepheid assays as previously reported for upper respiratory infections such as SC2 (17,18).

The study had a 4.4% rate of discrepant samples across all four analytes. Discrepant results between two platforms can occur due to a variety of reasons such as differences in specimen collection and differences in analytical sensitivity of the assays. Most of the discrepant results in the clinical study (∼75%) had cycle threshold values > 37 on the platform that called a positive indicating low viral loads in these samples. These samples could have been missed due to viral load being close to the analytical sensitivity of the platform. All the discrepant results for samples with co-infections can be categorized as low viral load samples (Ct > 37) for the discrepant result. Discrepant resolution using a reference method such as viral infectivity testing or genetic sequencing was not used (19).

Two ANS samples were collected from each participant in the clinical study. While good clinical practices were followed to switch the order of collection in each buffer and care was taken while collecting the samples, there was no way to control the amount of viral targets present at the time of sampling in each of a pair of swabs due to possible biological reasons related to viral shedding (20,21). Although the Fast PCR MRP test had good agreement with the comparator, sampling error could be a potential cause of discrepant results and could explain why most of the discrepant pairs had a Ct > 37, suggestive of a low viral load. However, the near perfect concordance between Fast PCR MRP and comparator (Cohen’s kappa > 0.88 for all assays) is suggestive that sampling error did not introduce significant bias in the outcome of the study.

In conclusion, this clinical study showed that the Fast PCR MRP test is a sensitive and specific assay to aid in the diagnosis of infections due to Flu A, Flu B, RSV and SC2. With its ease of use, sample-to-answer time of ∼10 minutes and cloud connectivity, the Fast PCR MRP System facilitates rapid diagnosis of these common respiratory infections.

## Data Availability

All raw data produced in the present study are available upon reasonable request to the authors. All processed data is contained in the manuscript.

## Acknowledgements

We acknowledge the staff at the participating clinical sites, site coordinators at MDC Associates, Karen Copeland at Boulder Statistics LLC, Binh Do and Gabriel Rodriguez for their help in shipping RUO kits to participating sites and sample management at AMDI, Corinne Minzer, Francisco Hoang and Kenzo Katsube for their help in manufacturing the RUO kits, and the AMDI Technical Support Team for handling phone calls from participating sites without whom the study would not have been possible.

We offer our special thanks to the study participants at all the clinical sites.

## Supplementary Information

**Table S1:** Inclusion/Exclusion Criteria for Prospectively Enrolled Participants.

| Inclusion Criteria | Exclusion Criteria |
| --- | --- |
| Subjects with one or more signs/symptoms of respiratory infection, including but not limited to fever, cough, sore throat, runny nose, myalgia, headache, chills or fatigue. | Subject or legal guardian is unable or unwilling to provide Informed Consent or assent (if required) |
| If ≥ age of majority, subject has provided Informed Consent.<br>If $<$ age of majority, parent/legal guardian consent provided, and subject assent provided.<br>The following consent/assent is required per age group:<br>- Minors $<7$ years of age: written consent by the parent /legal guardian.<br>- Minors 7 to the age of majority will provide assent (specific for age-appropriate language) followed by parent/legal guardian consent.<br>- Age of majority: written consent | Subject is unable or unwilling to provide the required specimens |
| Subject willing to provide 2 anterior nasal swab (ANS) specimens. | Subjects who have received an influenza vaccine by an influenza nasal spray/mist vaccine within the previous 30 days |
|  | Subject's healthcare provider determined that specimen collection represented an unacceptable healthcare risk |
|  | Previously participated in the study |

**Table S2:**
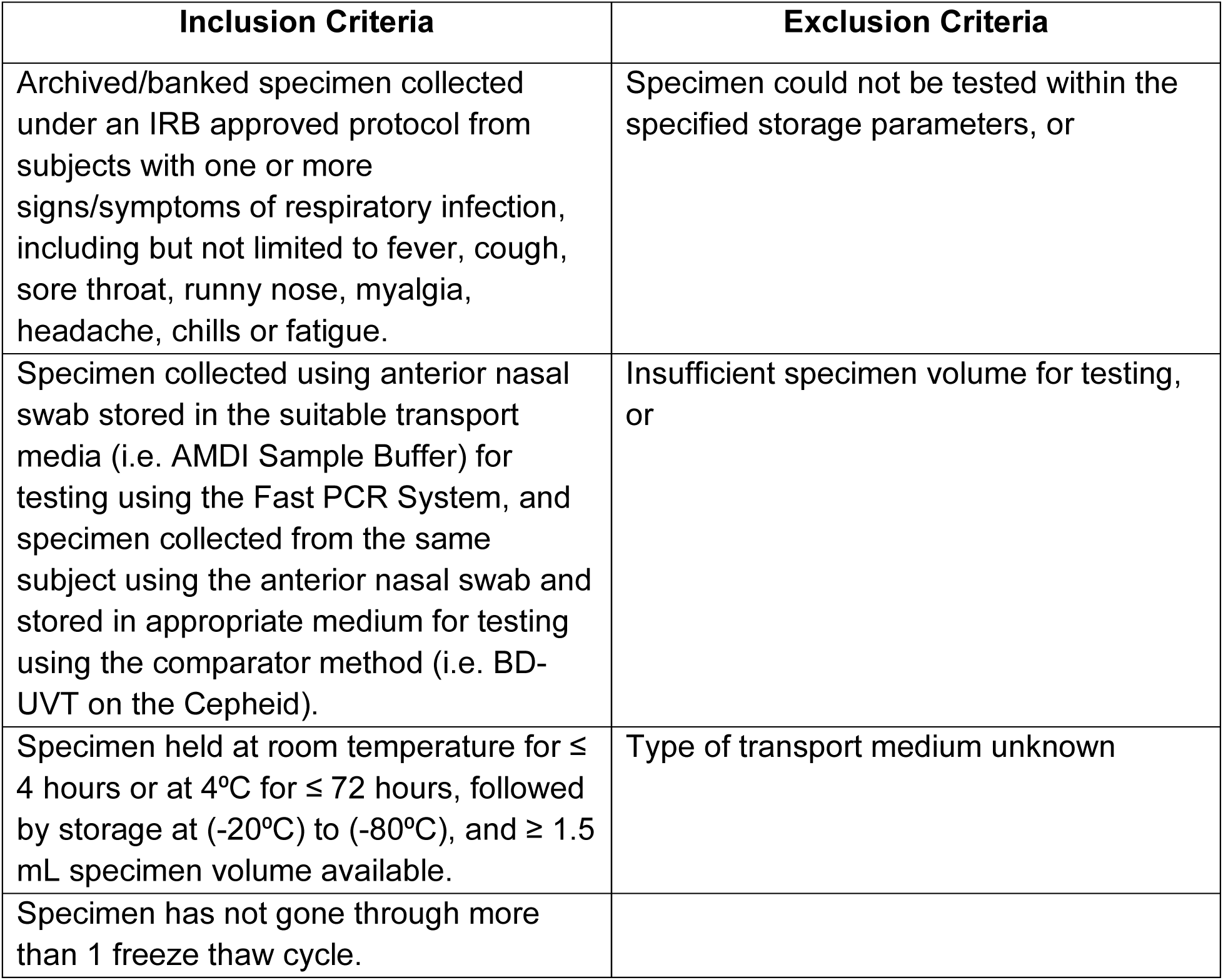
Inclusion/Exclusion Criteria for Archived Samples.

**Table S3:** Age Demographics in the Evaluable Participants.

| **Age Group** | **All**<br>**n (%)** | **Prospective**<br>**n (%)** | **Archived**<br>**n (%)** |
| --- | --- | --- | --- |
| All | 1,906 | 1,788 | 118 |
| $\leq 5$ yr | 98 (5.1%) | 94 (5.3%) | 4 (3.4%) |
| 6-18 yr | 344 (18.0%) | 311 (17.4%) | 33 (28.0%) |
| 19-40 yr | 687 (36.0%) | 664 (37.1%) | 23 (19.5%) |
| 41-60 yr | 519 (27.2%) | 496 (27.7%) | 23 (19.5%) |
| $\geq 61$ yr | 258 (13.5%) | 223 (12.5%) | 35 (29.7%) |

**Table S4:** Race Demographics in the Evaluable Participants.

| Race | All | Prospective | Archived |
| --- | --- | --- | --- |
|  | n (%) | n (%) | n (%) |
| All | 1,906 | 1,788 | 118 |
| Am. Indian or Alaskan Nat. | 4 (0.2%) | 4 (0.2%) | 0 (0.0%) |
| Asian | 86 (4.5%) | 81 (4.5%) | 5 (4.2%) |
| Black or African Am. | 284 (14.9%) | 283 (15.8%) | 1 (0.8%) |
| Not Reported | 31 (1.6%) | 29 (1.6%) | 2 (1.7%) |
| Other or Mixed Race | 70 (3.7%) | 70 (3.9%) | 0 (0.0%) |
| Unknown | 21 (1.1%) | 20 (1.1%) | 1 (0.8%) |
| White | 1410 (74.0%) | 1301 (72.8%) | 109 (92.4%) |

**Table S5:**
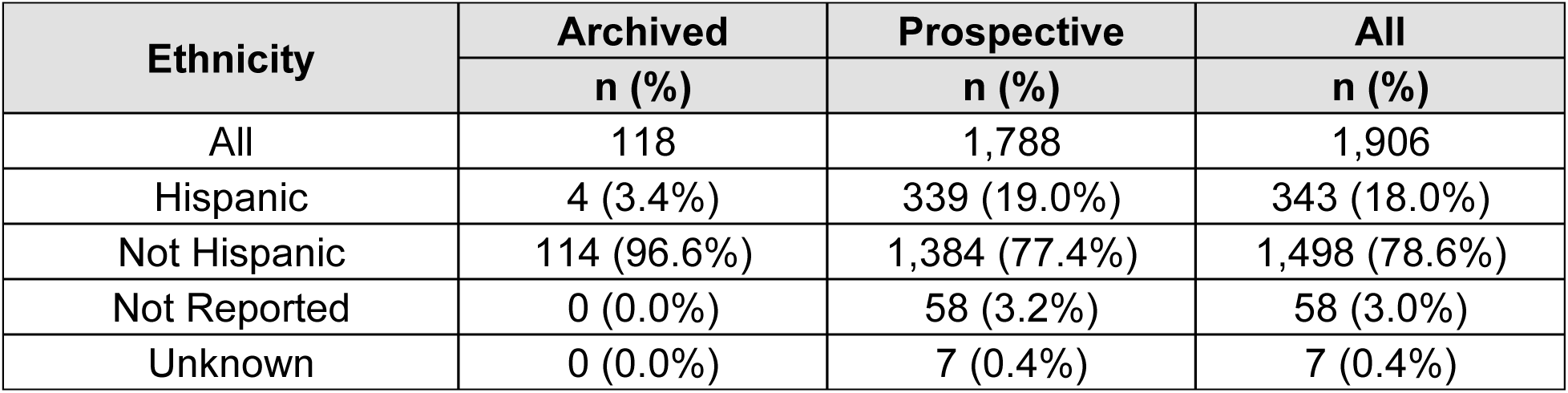
Ethnicity Demographics in the Evaluable Participants.

**Table S6:**
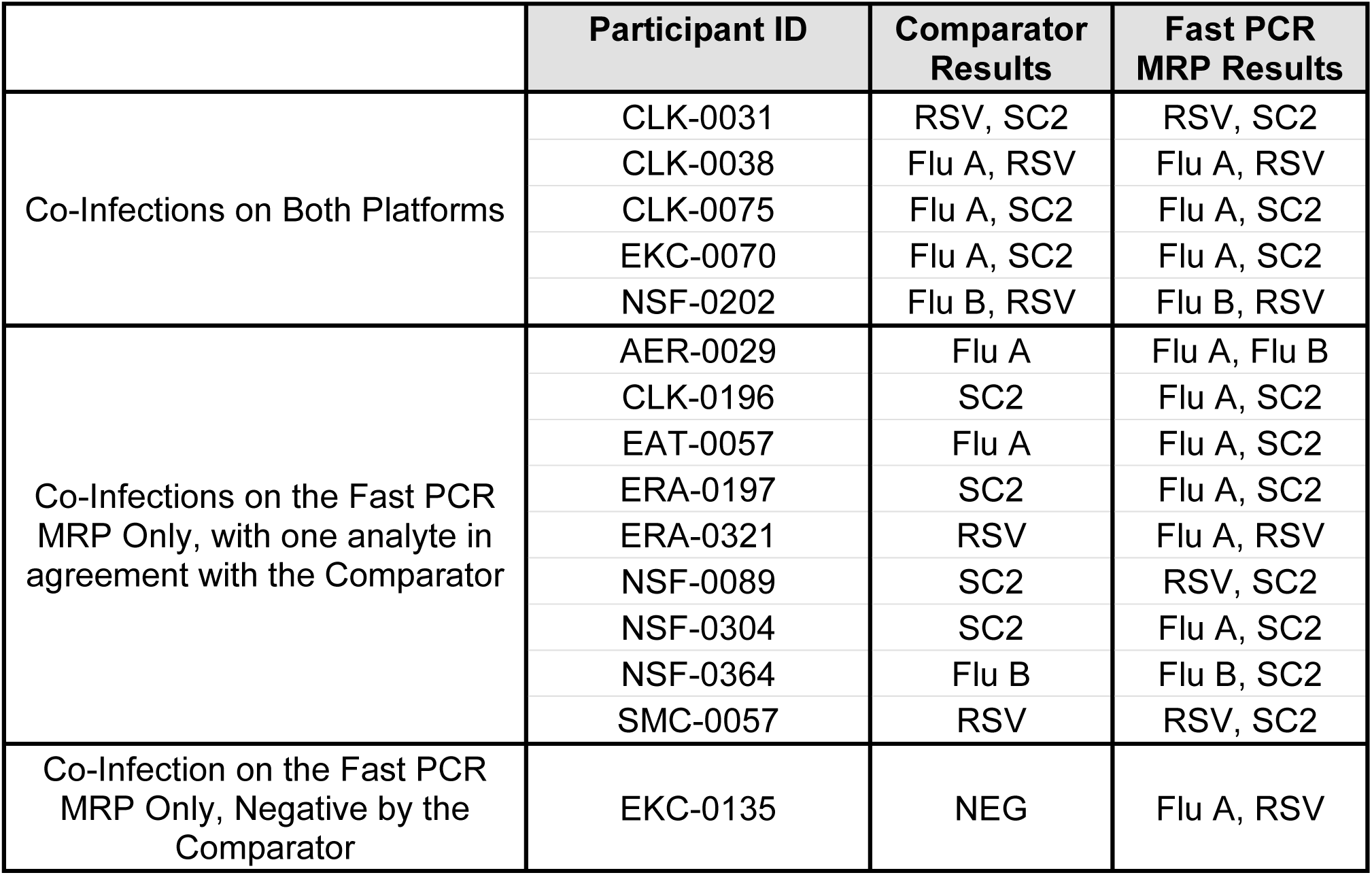
Clinical Samples with possible co-infections and results using Fast PCR MRP and Comparator tests.

